# Reproducibility of global electrical heterogeneity measurements on 12-lead ECG: The Multi-Ethnic Study of Atherosclerosis

**DOI:** 10.1101/2021.06.07.21258521

**Authors:** Kazi T. Haq, Katherine J. Lutz, Kyle Peters, Natalie Craig, Evan Mitchell, Anish K. Desai, Nathan W. L. Stencel, Elsayed Z. Soliman, João A.C. Lima, Larisa G. Tereshchenko

## Abstract

**Objective:** Vectorcardiographic (VCG) global electrical heterogeneity (GEH) metrics showed clinical usefulness. We aimed to assess the reproducibility of GEH metrics.

**Methods:** GEH was measured on two 10-second 12-lead ECGs recorded on the same day in 4,316 participants of the Multi-Ethnic Study of Atherosclerosis (age 69.4±9.4 y; 2317(54%) female, 1728 (40%) white, 1138(26%) African-American, 519(12%) Asian-American, 931(22%) Hispanic-American). GEH was measured on a median beat, comprised of the normal sinus (N), atrial fibrillation/flutter (S), and ventricular-paced (VP) beats. Spatial ventricular gradient’s (SVG’s) scalar was measured as sum absolute QRST integral (SAIQRST) and vector magnitude QT integral (VMQTi).

**Results:** Two N ECGs with heart rate (HR) bias of -0.64 (95% limits of agreement [LOA] - 5.68 to 5.21) showed spatial area QRS-T angle (aQRST) bias of -0.12 (95%LOA -14.8 to 14.5). Two S ECGs with HR bias of 0.20 (95%LOA -15.8 to 16.2) showed aQRST bias of 1.37 (95%LOA -33.2 to 35.9). Two VP ECGs with HR bias of 0.25 (95%LOA -3.0 to 3.5) showed aQRST bias of -1.03 (95%LOA -11.9 to 9.9). After excluding premature arial or ventricular beat and two additional beats (before and after extrasystole), the number of cardiac beats included in a median beat did not affect the GEH reproducibility. Mean-centered log-transformed values of SAIQRST and VMQTi demonstrated perfect agreement (Bias 0; 95%LOA -0.092 to 0.092).

**Conclusion:** GEH measurements on N, S, and VP median beats are reproducible. SVG’s scalar can be measured as either SAIQRST or VMQTi.

**Significance:** Satisfactory reproducibility of GEH metrics supports their implementation.

**Highlights:**

- VCG metrics are reliably reproducible, which supports their implementation.
- GEH is reproducible if measured during atrial fibrillation or ventricular pacing.
- Scalar of spatial ventricular gradient can be measured as either SAIQRST or VMQTi.

## 1. Introduction

Vectorcardiographic (VCG) global electrical heterogeneity (GEH) metrics showed their clinical usefulness in heart failure patients with implanted primary prevention cardioverter-defibrillators[1-4] and in the general population.[5-10] GEH metrics represent comprehensive characterization of spatial ventricular gradient (SVG) magnitude, direction (azimuth and elevation), spatial QRS-T angle, and SVG’s scalar value, sum absolute QRST integral (SAIQRST).[11, 12] Spatial QRS-T angle is a well-recognized and extensively studied marker of cardiovascular risk.[13] SVG defines a vector along which non-uniformity in excitation and repolarization is the most prominent.[14, 15] Wilson’s frontal plane ventricular gradient was extended into three-dimensional (3D) SVG in 1954.[16, 17] The GEH concept is based on a strong scientific premise and more than 80 years of theoretical, experimental, and clinical investigations. Extensive previous studies support the need to implement GEH in clinical practice.[1-12, 18-24] For successful implementation in clinical practice, it is necessary to assess the reproducibility of GEH metrics.

A small preliminary study of GEH reproducibility was previously conducted using 5-min recordings of high-resolution (1000Hz) ECG signals, comparing two randomly selected 10-second segments.[25] However, the reproducibility of GEH measured on routine clinical 10-second 12-lead ECG has not been studied. Moreover, it is unknown if there are differences in the reproducibility of GEH measured on the different types of median beat (normal sinus, atrial fibrillation, ventricular paced), and whether the number of beats included in a median beat and removal of a premature beat (either supraventricular or ventricular) affects reproducibility. To address these knowledge gaps, we conducted a comprehensive study of GEH reproducibility. We hypothesized that GEH measurements on 10-second 12-lead ECG are reproducible.

## 2. Material and Methods

### 2.1. Study population

We conducted an ancillary study in the Multi-Ethnic Study of Atherosclerosis (MESA).[26] All MESA participants signed informed consent before entering the MESA study. All MESA protocols were reviewed and approved by the local MESA field center IRBs. The current study protocol was approved by the Oregon Health & Science University (OHSU) institutional review board.

MESA study participants had routine resting 12-lead ECG recorded during the first (years 2000-2002) and the fifth (years 2010-2011) study examinations. We aimed to assess the reproducibility of both normal sinus (N) and abnormal types of median beats, including ventricular-paced (VP) and supraventricular (S, due to atrial fibrillation or atrial flutter) median beats. Therefore, we elected to analyze the data of the 5^th^ MESA examination that included older participants with a higher probability of having S and VP median beats. In the present study, we included MESA participants who had at least two 10-second resting 12-lead ECGs recorded consecutively during the 5^th^ examination. To be eligible for the reproducibility study, it was required to have at least two consecutive 10-second ECG recordings in the same rhythm (e.g., N and N, S and S, VP and VP). We excluded participants who did not have two consecutive 10-second ECG recordings available, had a median beat other than N, S, or VP types, or the rhythm (and thus the median beat) changed between 2 consecutive ECG recordings.

### 2.2. Theory, measurements, and calculation

Twelve-lead digital ECGs were obtained by trained technicians using GE MAC 1200 electrocardiographs with standardized procedures. ECGs were transmitted electronically to the MESA ECG Reading Center located at the Epidemiological Cardiology Research Center (Wake Forest School of Medicine, Winston-Salem, NC). According to MESA protocol, all filters in the ECG machines were disabled to provide unfiltered measurements. Initially, ECGs were automatically processed, after visual inspection for technical errors and inadequate quality, using the 2001 version of the GE Marquette 12-SL program.

For the purpose of this analysis, raw digital ECG signal was analyzed in the Tereshchenko laboratory at OHSU, as previously described.[7, 20, 27] Briefly, the analysis includes several steps. Each cardiac beat was manually labeled by at least two physician investigators (KJL, KP, EM, NC, AKD, NWLS, LGT). Then, 12-lead ECG was transformed into XYZ ECG, using a Kors transformation.[28] The origin of the heart vector was identified, and the time-coherent global median beat was constructed.[27] Only one (dominant) type of beat was included in the development of a median beat. This study included only three types of median beat: N, S, and VP. Ectopic beats (both atrial and ventricular premature beats) and two additional beats (the beat before and beat after ectopic beats) were excluded, and such fact was noted. We calculated the number of beats in a 10-second ECG recording and the number of beats included in the median beat.

Scalar values of SVG were measured by sum absolute QRST integral (SAIQRST) and by QT integral on vector magnitude (VM) signal (VMQTi).[20] Both area and peak QRS-T angles were measured.[7, 20, 27] Quality control of automated ECG analysis was performed by the investigator (KTH) with the aid of visual display. The open-source MATLAB (MathWorks, Natick, MA, USA) code is provided at https://physionet.org/physiotools/geh & https://github.com/Tereshchenkolab/Origin. Figure 1 illustrates the VCG measurements.

**Figure 1.**
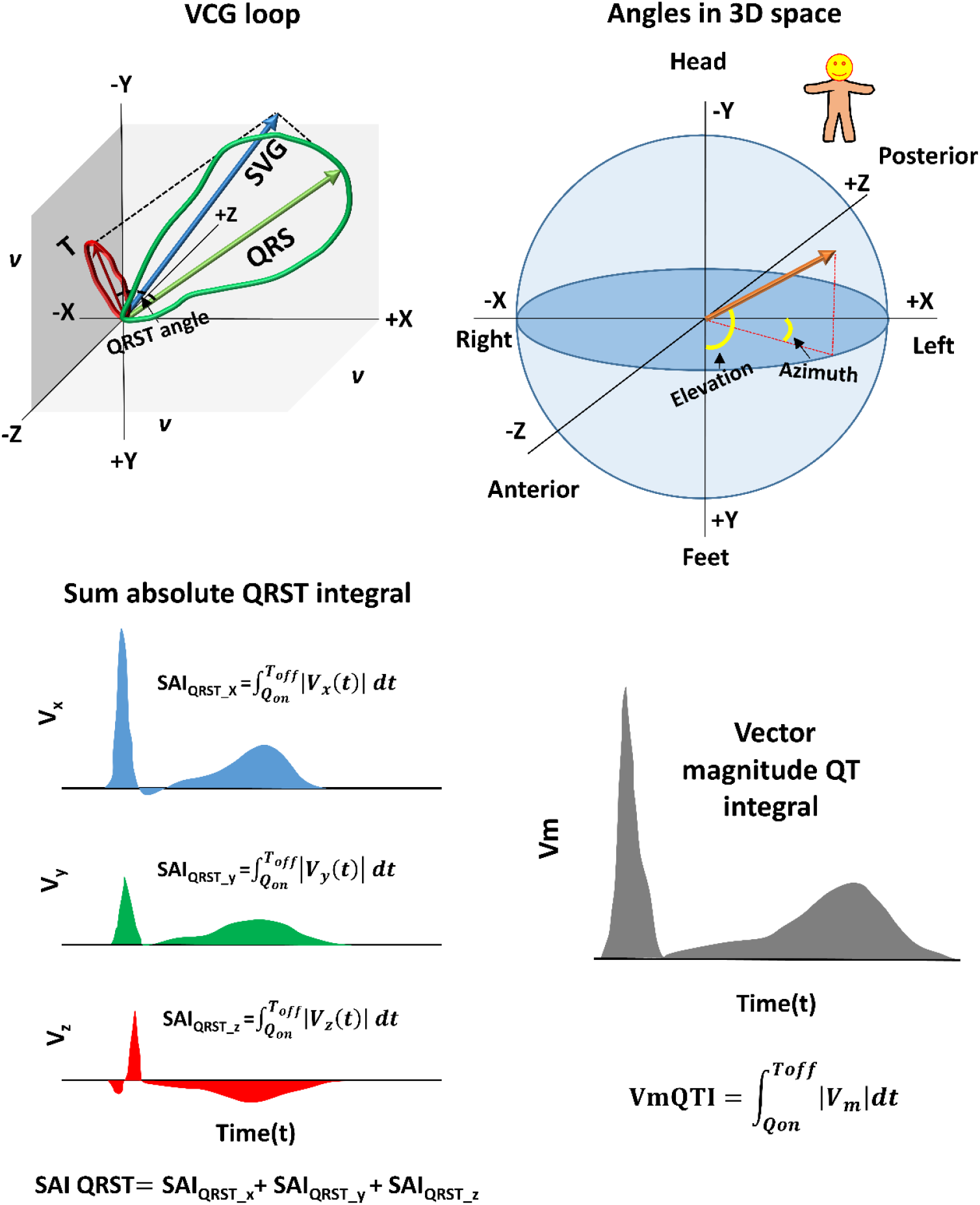
GEH measurements. (**A**) Spatial ventricular gradient (SVG) vector (blue) obtained as a vector sum of QRS (green) and T vector (red). Spatial QRS-T angle is the 3D angle between QRS and T vector. (**B**) Orientation of the angles (azimuth and elevation) in 3D space. (**C**) Sum absolute QRST integral (SAIQRST). (**D**) Vector magnitude QT integral.

Spatial peak and spatial area QRS, T, and spatial ventricular gradient (SVG) vectors were constructed, and their direction (azimuth and elevation) and magnitudes were measured.[7, 20, 27] Spatial peak QRS and T vectors connected origin point with the furthest points away from the origin point in the QRS-loop and T-loop, respectively.

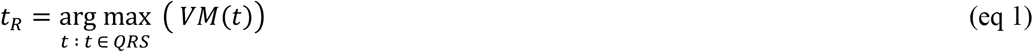

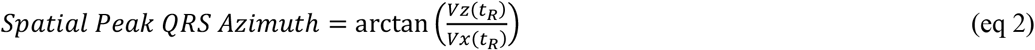

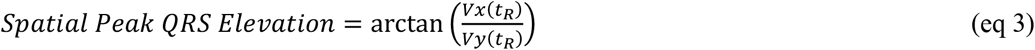

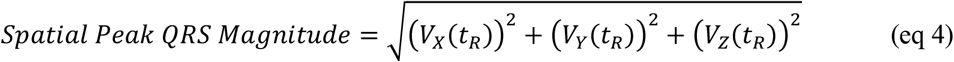

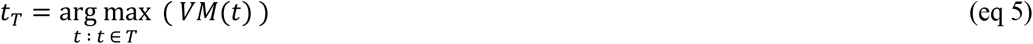

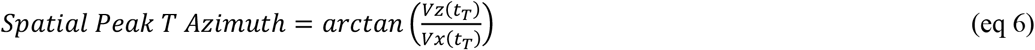

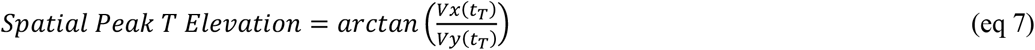

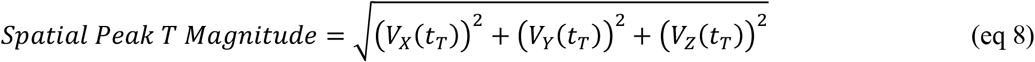

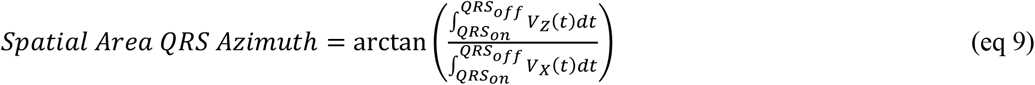

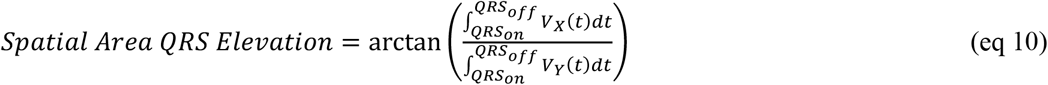

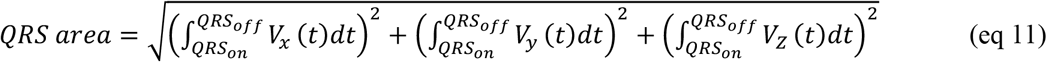

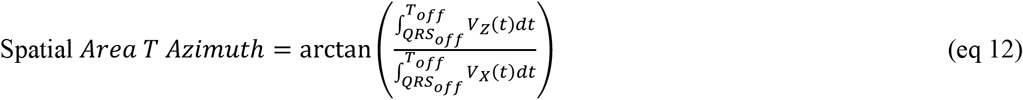

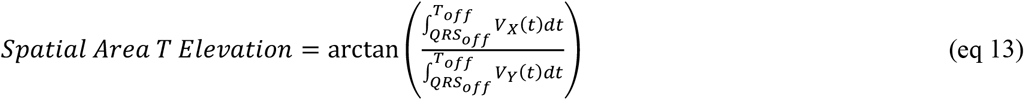

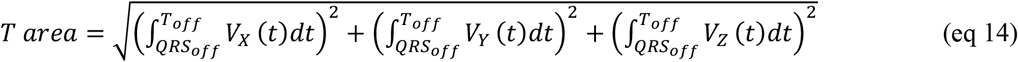

Magnitude and direction of the spatial area and peak SVG vectors were measured.

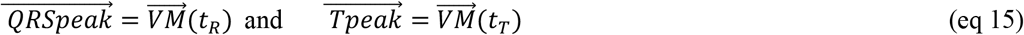

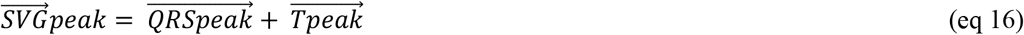

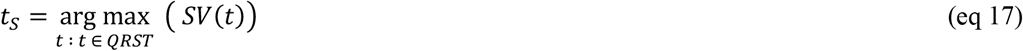

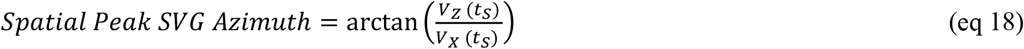

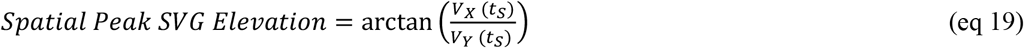

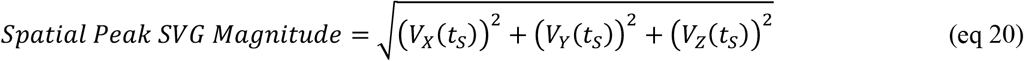

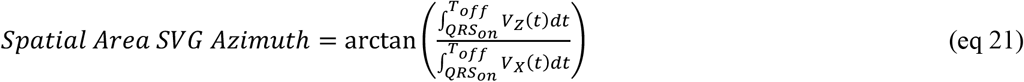

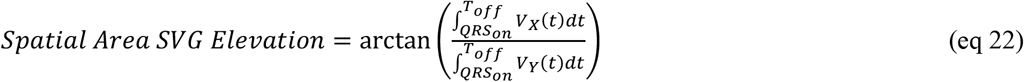

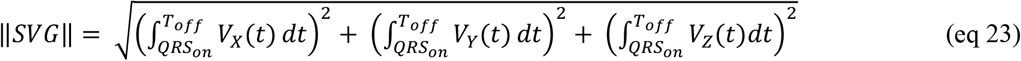

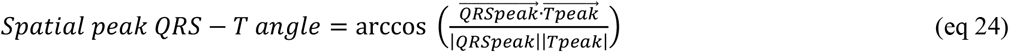

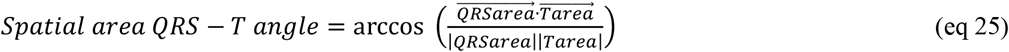

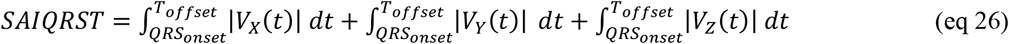

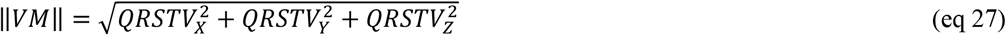

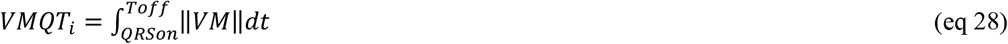

The reproducibility of the measurements on two consecutive 10-second ECGs was assessed by Bland-Altman analysis.[29, 30] The degree of the agreement was expressed as the bias (the mean difference) with 95% limits of agreement [mean±2 standard deviations(SD)], and relative bias (the mean difference of two measurements divided by their mean value). Precision was defined as 100% minus relative % bias.

Coefficient of variation (CV) was calculated as the ratio of the SD of residuals (RMSE) after fitting linear regression between ECG1 and ECG2 variables to the sample mean of the ECG1 variable, and then multiplied by 100 to express it in terms of a percentage. CV assesses the variability around the regression line relative to the mean of ECG1 variable.

Interclass correlation coefficient (ICC), equal to Cronbach’s alpha statistic,[31] was calculated for standardized variables (in the scale to mean 0 and variance 1).

The circular SVG azimuth variables range from -180° to +180°. Thus, to calculate relative bias, CV, and ICC, SVG azimuth variables were transformed by doubling their value and adding 360.

The statistical correlation between pairs for each parameter was calculated as Pearson’s correlation coefficient *r*. In addition, Lin’s concordance correlation coefficient ρ_c_ (rho_c) was calculated to describe the strength of agreement: >0.99 indicated almost perfect agreement; 0.95– 0.99, substantial agreement; 0.90–0.95, moderate agreement; <0.90, poor agreement.

Furthermore, the Bradley-Blackwood procedure was used to compare the means and variances of the 2 measurements simultaneously.[32]

To confirm that SAIQRST and VMQTi are essentially the same, we assessed the agreement between SAIQRST and VMQTi measured on the same ECG#1. We used mean-centered values of log-transformed SAIQRST and VMQTi, because of expected differences in their absolute values. The mean-centered value was obtained by subtracting the mean of the variable from each individual observation.

We compared the reproducibility of (1) three types of a median beat (N, S, VP), (2), and (4) on N median beat comprised of ≤ 6 beats, 7-9 beats, and ≥ 10 beats. Statistical analysis was conducted using STATA MP 16.1 (StataCorp LP, College Station, TX). Open-source STATA code was provided at https://github.com/Tereshchenkolab/statistics.

## 3. Results

### 3.1. Study population

The study flowchart is shown in Figure 2. After excluding the MESA study participants who did not meet the inclusion criteria, the study population included 4,316 participants.

**Figure 2.**
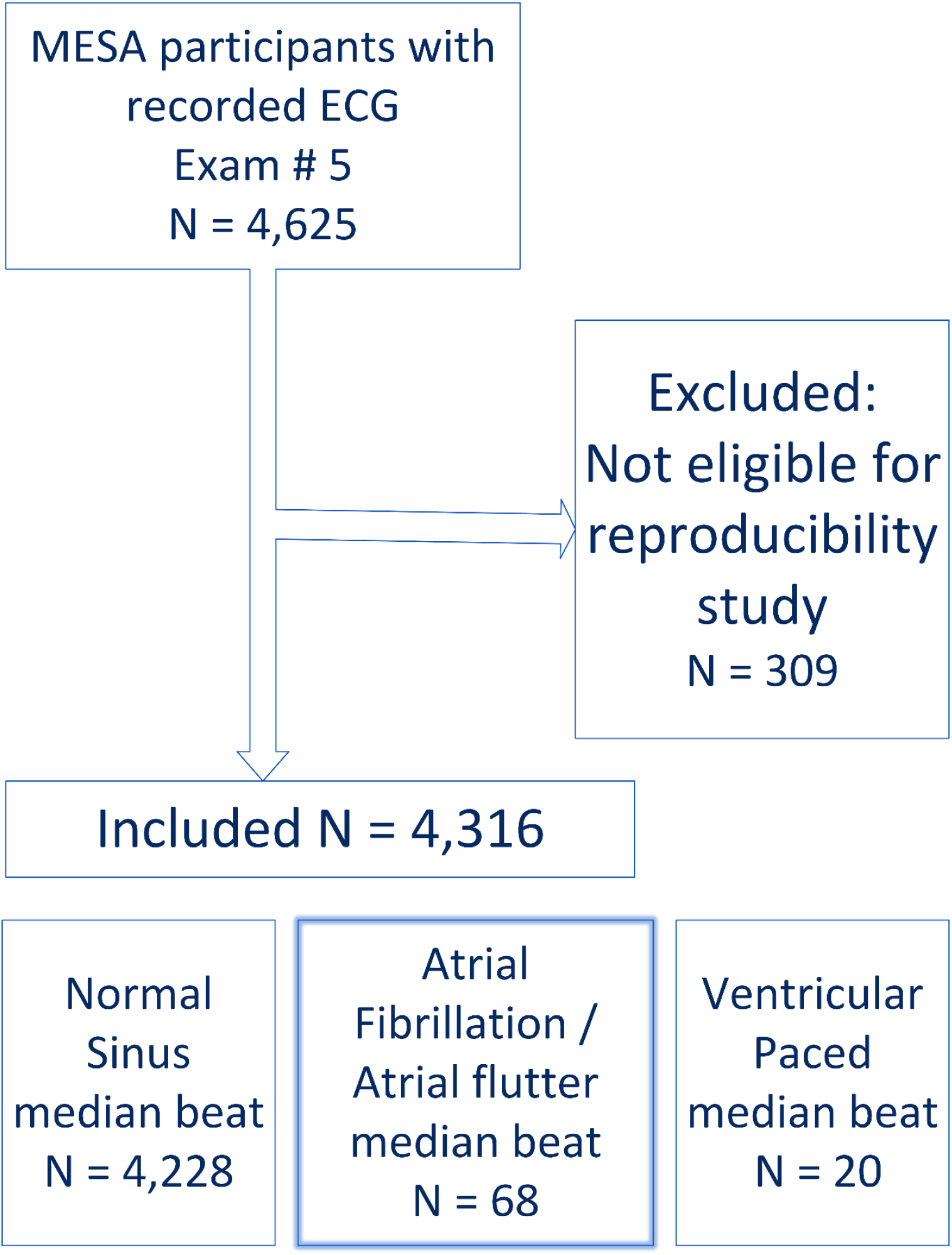
Study flowchart.

Demographic characteristics are shown in Table 1. The vast majority of participants had N median beats (n=4228; 98.0%), whereas only few had S median beats (n=68; 1.5%) and VP median beats (n=20; 0.5%). As expected, participants with N median beats were younger than those with S or VP median beats. Among participants with N median beats, 54% were female, and 40% were white.

**Table 1.** Demographic characteristics of the study participants.

| Characteristic | N beat (n=4228) | S beat (n=68) | VP beat (n=20) |
| --- | --- | --- | --- |
| Age (mean±SD), y | 69.2±9.3 | 77.5±8.3 | 76.3±10.5 |
| Women, n(%) | 2291(54) | 21(31) | 5(25) |
| White, n(%) | 1681(40) | 39(57) | 8(40) |
| African-American, n(%) | 1121(27) | 12(18) | 5(25) |
| Asian-American, n(%) | 517(12) | 2(3) | 0 |
| Hispanic-American, n(%) | 909(22) | 15(22) | 7(35) |

### 3.2. Reproducibility of GEH

Reproducibility of the GEH metrics measured on two consecutive 10-second ECGs on a normal sinus median beat ranged between perfect and substantial (Table 2), and relative bias was less than 1% for all GEH metrics. GEH measured on VP and S median beats was slightly less reproducible, ranging between substantial and moderate, and the relative bias was less than 5% for all GEH metrics. For most VCG metrics, 95% limits of agreement were similar for N and VP median beats (Figure 3) but were more prominent for S median beats.

**Table 2.**
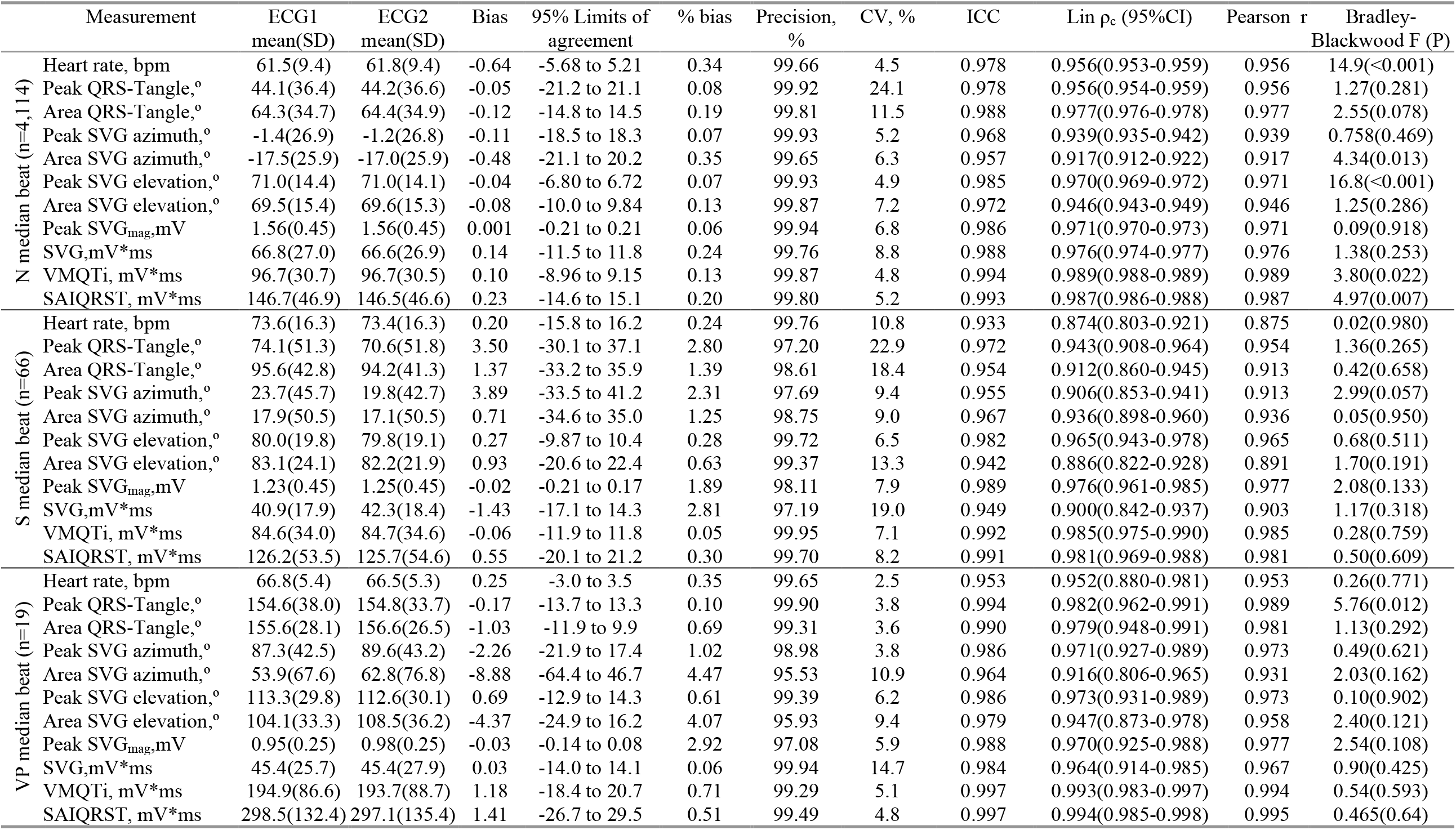
Reproducibility agreement of GEH measurements on two 10-second ECG recordings, on N, S, and VP median beats.

**Figure 3.**
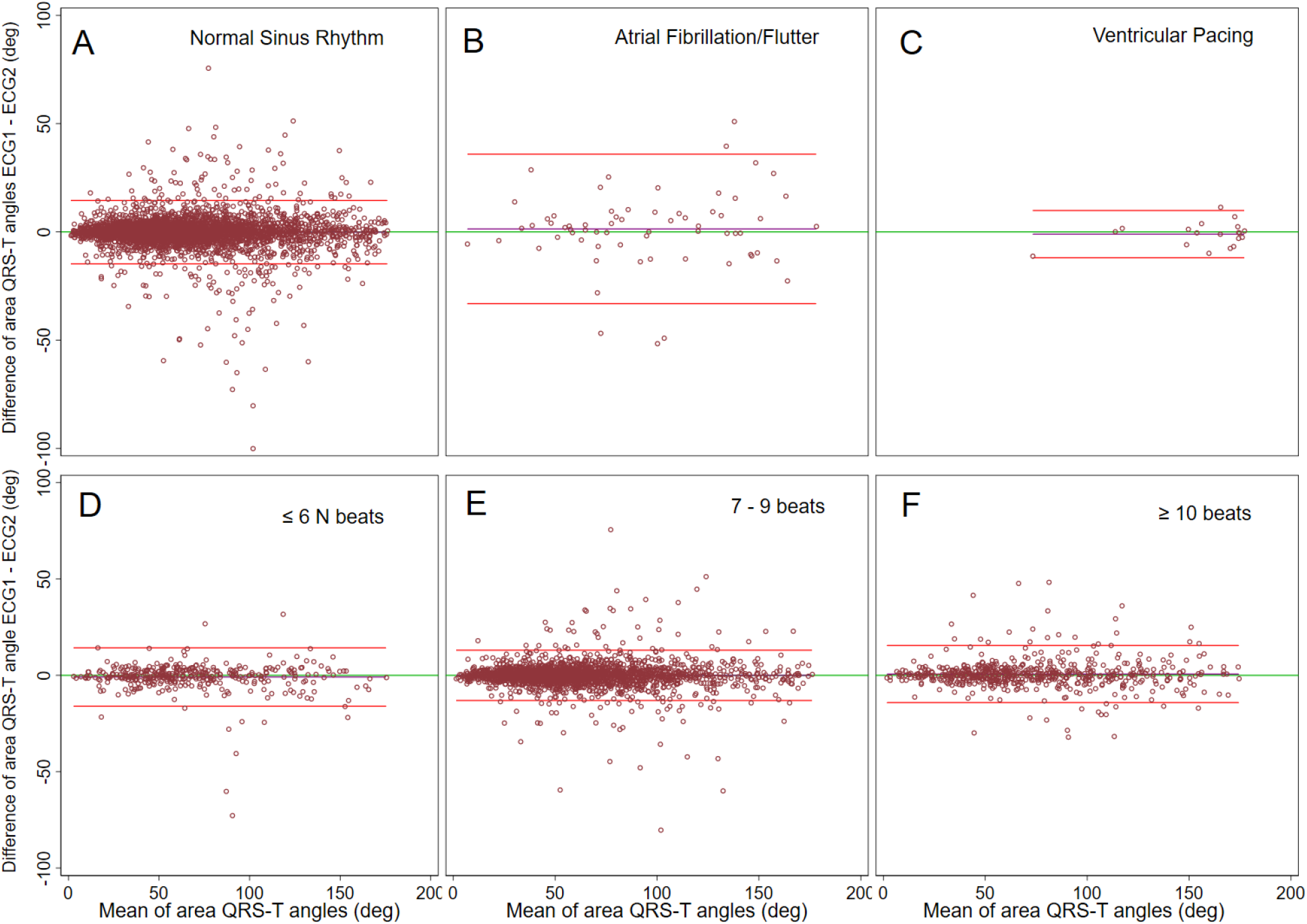
Bland-Altman plots demonstrating agreement of spatial area QRS-T angle on two ECGs. Median beats were constructed in (**A**) normal sinus (N) rhythm, (**B**) atrial fibrillation/flutter, (**C**) ventricular pacing, including (**D**) ≤6 N beats, (**E**) 7-9 N beats, (**F**) ≥10 N beats. The scatterplot presents paired differences (Y-axis), plotted against pair-wise means (X-axis). The reference line indicates the perfect average agreement, Y = 0. The central green line indicates the mean difference between the two measurements, or mean bias. Upper and lower lines represent the mean ± 2 standard deviations (SD), or 95% limits of agreement.

Complete exclusion of ECG recordings with previously excluded PVCs or PACs did not have a noticeable effect on the reproducibility of GEH metrics (Table 2). Overall, 95% limits of agreement were similar for all N median beats, regardless of the number of beats included in the template (Figure 3 and Table 4). There were no differences in GEH reproducibility in sex and race/ethnicity subgroups (data not shown).

**Table 3.** Reproducibility agreement of GEH measurements on two 10-sec ECGs on N beat without excluded premature beats.

| | Measurement | ECG1<br>mean(SD) | ECG2<br>mean(SD) | Bias | 95% Limits of<br>agreement | % bias | Precision,<br>% | CV,% | ICC | Lin $\rho_c$ (95%CI) | Pearson r | Bradley-Blackwood F (P) |
| --- | --- | --- | --- | --- | --- | --- | --- | --- | --- | --- | --- | --- |
| 100% N median beat<br>(n=3,953) | Heart rate, bpm | 61.5(9.4) | 61.7(9.4) | -0.23 | -5.5 to 5.0 | 0.34 | 99.66 | 4.3 | 0.979 | 0.958(0.956-0.961) | 0.959 | 15.0(<0.0001) |
|  | Peak QRS-Tangle,° | 43.7(36.0) | 43.8(36.2) | -0.05 | -21.1 to 21.0 | 0.08 | 99.92 | 24.2 | 0.977 | 0.956(0.953-0.958) | 0.956 | 1.26(0.285) |
|  | Area QRS-Tangle,° | 63.9(34.5) | 64.0(34.7) | -0.10 | -14.6 to 14.4 | 0.15 | 99.85 | 11.4 | 0.989 | 0.977(0.976-0.979) | 0.977 | 2.09(0.124) |
|  | Peak SVG azimuth,° | -1.4(26.9) | -1.3(26.7) | -0.13 | -18.5 to 18.3 | 0.08 | 99.92 | 5.2 | 0.968 | 0.939(0.935-0.942) | 0.939 | 0.91(0.403) |
|  | Area SVG azimuth,° | -17.5(25.7) | -17.0(25.8) | -0.53 | -21.4 to 20.4 | 0.38 | 99.62 | 6.4 | 0.955 | 0.914(0.909-0.919) | 0.914 | 5.26(0.005) |
|  | Peak SVG elevation,° | 71.0(14.4) | 71.0(14.1) | -0.04 | -6.8 to 6.8 | 0.07 | 99.93 | 4.9 | 0.985 | 0.970(0.968-0.972) | 0.971 | 16.6(<0.0001) |
|  | Area SVG elevation,° | 69.5(15.3) | 69.6(15.2) | -0.08 | -9.8 to 9.6 | 0.14 | 99.86 | 7.1 | 0.973 | 0.947(0.944-0.951) | 0.947 | 1.57(0.209) |
|  | Peak SVG<sub>mag</sub>,mV | 1.57(0.45) | 1.56(0.45) | 0.001 | -0.21 to 0.21 | 0.06 | 99.94 | 6.8 | 0.985 | 0.971(0.969-0.973) | 0.971 | 0.08(0.926) |
|  | SVG,mV*ms | 67.0(27.1) | 66.9(27.0) | 0.15 | -11.4 to 11.7 | 0.27 | 99.73 | 8.8 | 0.988 | 0.976(0.975-0.978) | 0.976 | 1.79(0.167) |
|  | VMQT<sub>i</sub>, mV*ms | 96.8(30.5) | 96.7(30.3) | 0.10 | -8.87 to 9.08 | 0.13 | 99.87 | 4.7 | 0.994 | 0.989(0.988-0.989) | 0.989 | 4.10(0.017) |
|  | SAIQRST, mV*ms | 146.7(46.6) | 146.5(46.3) | 0.251 | -14.5 to 15.0 | 0.22 | 99.78 | 5.1 | 0.993 | 0.987(0.986-0.988) | 0.987 | 5.67(0.003) |

**Table 4.**
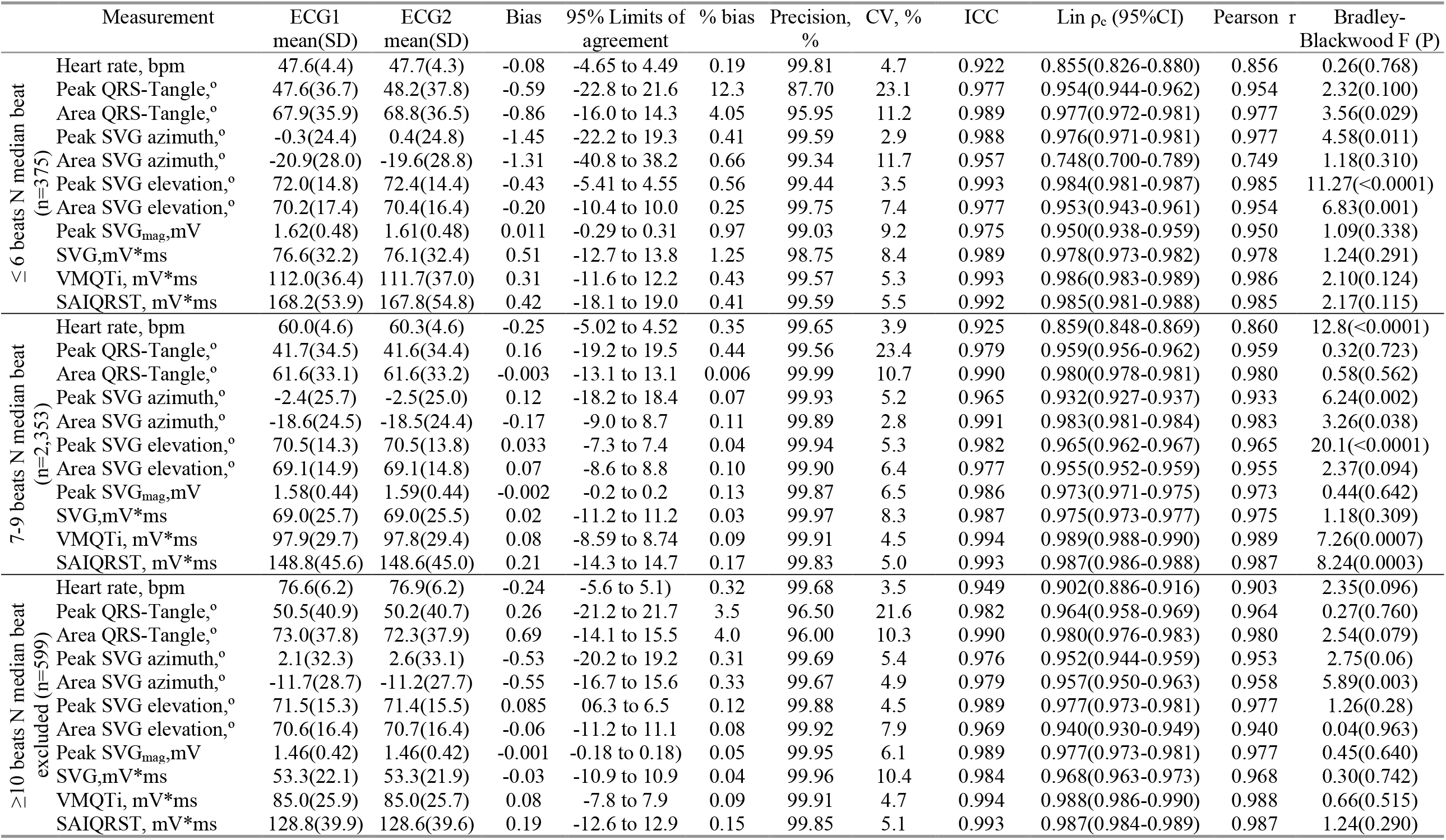
Reproducibility agreement of GEH metrics on two 10-sec ECGs, if ≤6, 7-9, ≥10 N beats included in N median beat.

### 3.3. Agreement between SAIQRST and VMQTi

Mean-centered log-transformed values of SAIQRST and VMQTi measured on the same ECG demonstrated perfect agreement. Bias was equal to zero with 95% limits of agreement from -0.092 to 0.092 (Figure 4).

**Figure 4.**
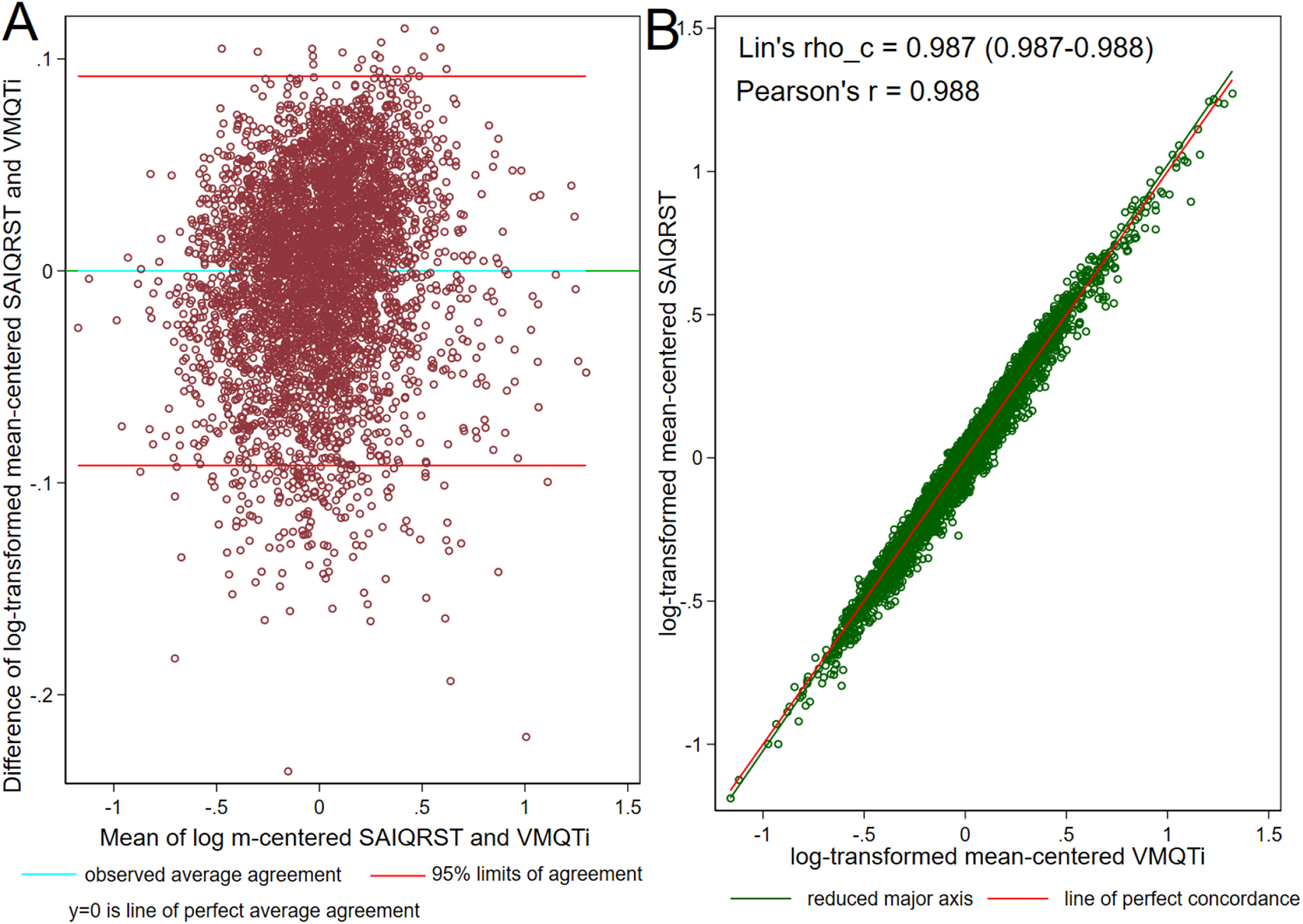
(**A**). Bland-Altman plot demonstrating agreement of mean-centered log-transformed SAIQRST and VMQTi. Definitions given in Figure 3 legend. (**B**). Concordance scatterplot of the mean-centered log-transformed SAIQRST and VMQTi. The reduced major axis of the data (green) goes through the intersection of the means and has the slope given by the sign of Pearson’s *r* and the ratio of the standard deviations. The reference red line shows the perfect concordance, Y = X.

## 4. Discussion

In this large study of more than 4,000 MESA participants with two consecutive routine clinical 12-lead ECG recordings, we observed high reproducibility of VCG metrics measured on several types of median beat: N, S, and VP. This is an important finding, confirming that GEH can be reproducibly measured not only during normal sinus rhythm but also during atrial fibrillation, atrial flutter, or ventricular pacing. After exclusion of either atrial or ventricular extrasystole and two additional beats (beats before and after an ectopic beat), the number of normal sinus cardiac beats included in the construction of N median beat did not affect the reproducibility of GEH metrics. In addition, this study confirmed that SVG’s scalar metric could be measured as either SAIQRST or VMQTi, as mean-centered log-transformed values of SAIQRST and VMQTi are nearly identical.

Satisfactory reproducibility of a biomarker is an important requirement for its clinical and research use. The biomedical research community set off alarm bells about the reproducibility crisis.[33] In response, our study assessed the reproducibility of GEH measured on two 10-second ECG recordings, mimicking a common clinical scenario. Knowledge about 95% limits of agreement is essential for the appropriate interpretation of any given measurement. Importantly, as ECG metrics are heart rate–dependent, the reproducibility of ECG metrics should therefore be considered within the context of heart rate. For example, we observed that if a heart rate on two ECG differed by 5-6 bpm, area QRS-T angle differed by 15 degrees. If a heart rate on 2 ECGs differed by ∼16 bpm (e.g., atrial fibrillation), area QRS-T angle differed by ∼36 degrees. Both measurement error and physiological effect of heart rate differences contribute to the degree of agreement between ECG measurements on two 10-second ECGs.

Only few and mostly small previous studies assessed the reproducibility of ECG biomarkers.[34-39] The earlier small (n=253) GEH reproducibility study included primarily African Americans, performed only N median beat analysis, and demonstrated similar findings to the current study.[25] In two random 10-second ECG segments selected in 5-minute ECG recording, a heart rate differed by ∼5 bpm, and spatial QRS-T angle differed by ∼13 degrees. Notably, in the current study, we reported GEH reproducibility on abnormal median beats (S and VP) for the first time.

A previous study of the reproducibility of automated 12-lead ECG measurement[30] showed that the agreement for QRS duration measured on a global median beat (95% limits of agreement ±9 ms) was better than for QRS duration measured on a median beat of each individual ECG lead (95% limits of agreement ∼13 ms). Interestingly, relative bias was especially high (∼3%) for T wave amplitude measured on individual leads V1-V4 on an N median beat.[30] In contrast, in the current study, the relative bias for VCG magnitudes measured on a global time-coherent N median beat was less than 1%, supporting the importance of an appropriate VCG’s origin point detection.[27]

Young et al. [40] conducted a simulation study and investigated the effect of inaccuracies in QRS_onset_, QRS_offset_, and T_offset_ detection on spatial QRS-T angle. They observed the mean absolute error up to ∼ 30 degrees, which was slightly larger than the 95% limits of agreement observed in the current study (up to ±22 degrees). Furthermore, it is important to consider differences in signal processing approach between the two studies: Young et al. [40] (1) applied a 0.5 – 45 Hz bandpass filter that can modulate QRST morphology, (2) excluded from analysis ECGs with small T-wave amplitudes (if maximum T/QRS < 0.1), and (3) defined VCG’s origin point .at the beginning of QRS complex. We agree that the approach of Young et al. [40] is a reasonable approach for fully automated measurements. However, physiologically, PR interval reflects atrial repolarization, which is an electrically active phase frequently responsible for heart vector deviation and does not meet the definition of electrical quiescence. Moreover, the filter’s low-pass band of 45Hz significantly affects the amplitudes and morphology of the QRS complex.

In addition, this study confirmed that either SAIQRST or VMQTi could be used to quantify SVG’s scalar, as the mean-centered log-transformed values of these two variables were nearly identical. Relative bias and 95% limits of agreement were smaller for VMQTi than SAIQRST, suggesting that the reproducibility of VMQTi is slightly superior than SAIQRST. Only two fiducial points (QRS_onset_ and T_offset_) are required for VMQTi measurement, whereas six fiducial points (QRS_onset_ and T_offset_ on 3 XYZ leads) are required for SAIQRST measurements, thus increasing the probability of error.

## 5. Conclusions

VCG GEH measurements on a 10-second resting clinical ECG in participants with normal sinus rhythm, atrial fibrillation/flutter, or ventricular pacing are reproducible. After excluding a premature ectopic beat and two additional beats (before and after extrasystole), the number of cardiac beats included in a median beat template does not affect the reproducibility of VCG GEH measurements. The high reproducibility of GEH measurements supports their implementation in clinical practice and research.

## Data Availability

The MESA Study data are available through the National Heart, Lung, and Blood Institute's Biological Specimen and Data Repository Information Coordinating Center (BioLINCC), the National Center of Biotechnology Information's database of Genotypes and Phenotypes (dbGaP), and via MESA Coordinating Center at the University of Washington.
The open-source MATLAB (MathWorks, Natick, MA, USA) code is provided at https://physionet.org/physiotools/geh & https://github.com/Tereshchenkolab/Origin. 
Open-source STATA code was provided at https://github.com/Tereshchenkolab/statistics.

https://github.com/Tereshchenkolab

## Acknowledgments

This research was supported by contracts 75N92020D00001, HHSN268201500003I, N01-HC-95159, 75N92020D00005, N01-HC-95160, 75N92020D00002, N01-HC-95161, 75N92020D00003, N01-HC-95162, 75N92020D00006, N01-HC-95163, 75N92020D00004, N01-HC-95164, 75N92020D00007, N01-HC-95165, N01-HC-95166, N01-HC-95167, N01-HC-95168 and N01-HC-95169 from the National Heart, Lung, and Blood Institute, and by grants UL1-TR-000040, UL1-TR-001079, and UL1-TR-001420 from the National Center for Advancing Translational Sciences (NCATS). The authors thank the other investigators, the staff, and the participants of the MESA study for their valuable contributions. A full list of participating MESA investigators and institutions can be found at http://www.mesa-nhlbi.org.

